# COVID-19 vaccinations: perceptions and behaviours in people with primary ciliary dyskinesia

**DOI:** 10.1101/2021.11.08.21266047

**Authors:** Eva SL Pedersen, Christina M Mallet, Yin Ting Lam, Sara Bellu, Isabelle Cizeau, Fiona Copeland, Trini Lopez Fernandez, Michelle Manion, Amanda Harris, Jane S Lucas, Francesca Santamaria, COVID-PCD patient advisory group, Myrofora Goutaki, Claudia E Kuehni

**Affiliations:** Institute of Social and Preventive Medicine, University of Bern, Bern, Switzerland; Graduate School for Health Sciences, University of Bern, Switzerland; Associazione italiana Discinesia Ciliare Primaria Sindrome di Kartagener Onlus, Italy; Association ADCP, Saint-Étienne, France; PCD support UK, London, UK; Asociación Española de Pacientes con Discinesia Ciliar Primaria, Spain; PCD Foundation, United States; Primary Ciliary Dyskinesia Centre, NIHR Biomedical Research Centre, University Hospital Southampton NHS Foundation Trust, Southampton, UK; University of Southampton Faculty of Medicine, School of Clinical and Experimental Medicine, Southampton, UK; Department of Translational Medical Sciences, Federico II University, Naples, Italy; Division of Paediatric Respiratory Medicine and Allergology, Department of Paediatrics, Inselspital, Bern University Hospital, University of Bern, Switzerland; Associazione italiana Discinesia Ciliare Primaria Sindrome di Kartagener Onlus Italy; Association ADCP, France; PCD support UK; Selbsthilfegruppe Primäre Ciliäre Dyskinesie, Switzerland; PCD Australia Primary Ciliary Dyskinesia, Australia; PCD Foundation, USA; Verein Kartagener Syndrom und Primäre Ciliäre Dyskinesie, Germany

**Author notes:** **Corresponding author**: Prof. Claudia Kuehni, Institute of Social and Preventive Medicine, University of Bern, Mittelstrasse 43, 3012 Bern, Switzerland. (E.P), (C.M.), (Y.L), (M.G.), (C.K.).

**Keywords:** sars-cov-2, covid-19, pcd, primary ciliary dyskinesia, vaccine, vaccinations, pandemic

## Abstract

Primary ciliary dyskinesia (PCD) is a rare genetic disease that causes recurrent respiratory infections. People with PCD may be at high risk of severe COVID-19 and vaccination against SARS-CoV-2 is therefore important. We studied vaccination willingness, speed of vaccination uptake, side effects, and changes in social contact behavior after vaccination in people with PCD. We used data from COVID-PCD, an international participatory cohort study. A questionnaire was e-mailed to participants in May 2021 that asked about COVID-19 vaccinations. 423 participants from 31 countries replied (median age: 30 years; 261 (62%) female). Vaccination uptake and willingness was high with 273 of 287 adults (96%) being vaccinated or willing to be in June 2021; only 4% were hesitant. The most common reasons for hesitancy were fear of side effects (reported by 88%). Mild side effects were common but no participant reported severe side effects. Half of participants changed their social contact behaviour after vaccination by seeing friends and family more often. The high vaccination willingness in the study population might reflect the extraordinary effort taken by PCD support groups to inform people about COVID-19 vaccination. Clear and specific public information and involvement of representatives is important for high vaccine uptake.

## Introduction

Vaccination against COVID-19 has proven effective in preventing transmission of SARS-CoV-2 (1, 2) and most countries are vaccinating against COVID-19 (3). The first vaccines were administered at the end of 2020 and priority was given to people considered at high risk of severe COVID-19 such as elderly or people with chronic diseases (4). By August 2021, many European countries had vaccinated most of the people willing to get vaccinated. Vaccination willingness has generally been high but an important proportion of the general population in several countries still hesitate to get vaccinated (5-7). In some countries vaccine uptake has only reached 50% of the population (8).

There are little data about vaccination hesitancy among people considered at high risk of severe COVID-19. Primary ciliary dyskinesia (PCD) is a rare genetic multi-system disease where dysfunctional cilia lead to impaired mucociliary clearance, laterality defects, and other health problems (9-13). People with PCD have chronic upper and lower airway disease (9, 14-16), lung function is reduced, and some need supplementary oxygen (17-22). At the start of the pandemic, people with PCD and other chronic respiratory diseases were considered at high risk of severe disease, and people with PCD were therefore recommended to get vaccinated against SARS-CoV-2. To date, there is no information about vaccination willingness, vaccination uptake, and side effects among people with PCD. Studies including people with other pre-existing health conditions show varying vaccination willingness (23, 24). The speed of vaccination rollout differed between countries and it is unclear how fast high-risk populations were vaccinated. The aim of this study was to describe COVID-19 vaccination willingness and hesitancy among people with PCD, to study speed of vaccination uptake, assess reported side effects of the vaccines, and investigate changes in social contact behaviour and shielding after vaccination.

## Materials and methods

### Study design and inclusion criteria

We used data from COVID-PCD, an international cohort study that uses anonymous online questionnaires to collect information from people with PCD during the COVID-19 pandemic (clinicaltrials.gov: NCT04602481). COVID-PCD is a participatory research project where people with PCD have an active role in all stages of research from the design of the study, its content, the piloting, and communication of results. Details about the study methods and first results have been published (25, 26). In short, COVID-PCD includes persons of any age from anywhere in the world with a confirmed or suspected diagnosis of PCD. The study is designed for three age groups; children below 14 years, adolescents between 14 and 17 years, and adults aged 18 years or more. For children, the questionnaires are addressed to the parents, but the child is encouraged to help complete the questionnaires. Adolescents and adults complete the questionnaires themselves. The study is available in English, German, Spanish, Italian, and French and recruitment started on May 31, 2020. The Cantonal Ethics Committee of Bern approved the study (Study ID: 2020-00830). Informed consent to participate is provided online at the time of registration into the study. This article follows the STROBE reporting recommendations (27).

### Study procedures

The COVID-PCD study is conducted online. Participants are invited by PCD support groups who contact and inform people living with PCD through social media and email networks and encourage them to take part. The website (www.covid19pcd.ispm.ch) includes detailed information about the study and allows participants to register and consent via a link that leads them directly to the study database. Participants first complete a baseline questionnaire with questions on their disease, their usual symptoms, and SARS-CoV-2 infections experienced prior to joining the study (28). Thereafter they receive weekly follow-up questionnaires with questions on incident SARS-CoV-2 infections, COVID-19 vaccinations, current symptoms, and social contact behaviour. Intermittently, we send short special questionnaires that focus on specific topics. This paper presents data from a special questionnaire on vaccinations that was sent to participants on May 29^th^ 2021 and data on vaccinations retrieved through the weekly questionnaire. Participants received up to two reminders if they did not respond to the special questionnaire. Participating PCD support groups were strongly involved in the development of the questionnaires. All data were entered in a Research Electronic Data Capture (REDCap) database developed at Vanderbilt University (www.project-redcap.org) (29), which is securely hosted by the Swiss medical registries and data linkage centre (SwissRDL) at the University of Bern, Switzerland.

### Information about COVID-19 vaccinations

The special questionnaire on COVID-19 vaccinations asked whether participants had already been vaccinated and if yes which vaccine they received (supplementary table 1). We also asked whether participants changed their social contact behaviour after vaccination. Unvaccinated participants were asked about vaccination willingness. We distinguished between people who planned to get vaccinated, were not sure whether to get vaccinated, or did not want to get vaccinated. We further asked about reasons for and against vaccination.

The weekly questionnaire asked whether participants had been vaccinated since they completed the last weekly questionnaire, which date they were vaccinated, whether it was the first or the second shot, and which side effects they experienced. We asked about side effects separately after the first and the second vaccine shot.

### Statistical analyses

We described demographics of the participants, vaccination uptake, vaccination willingness and hesitancy, reasons for and against getting vaccinated, side effects, and change in social behaviour using number and proportion for categorical variables and mean and standard deviation (SD) or median and interquartile range for continuous variables. We described vaccination willingness and hesitancy in adults aged 18 years or above, adolescents aged between 12 and 17 years, and children aged 11 years or less. We chose the cut-off of 12 years because several vaccines had been approved for children down to the age of 12 years by June 2021. We described speed of vaccination using date of first vaccination from the weekly follow-up questionnaires and compared how fast participants living in different countries were vaccinated. To study factors associated with reported side effects, we used multilevel logistic regression. We ran a regression with each side effect as dependent variable (no side effects, local pain or swelling around injection site, tiredness, fever, muscle- or joint pain, other) and added the following explanatory variables: age, sex, type of vaccine, and timing of vaccine (first or second injection, included as a separate level representing repeated measurements within an individual). Our dataset had few missing values (less than 2% in single variables) and records with missing data were excluded from the analysis. We used STATA version 15 for statistical analysis.

## Results

423 of the 689 people (61%) who participated in the COVID-PCD study in May 2021 returned the special questionnaire. Median age was 30 years (age range: 1-85 years, interquartile range 12 to 47) and 261 (62%) were female (supplementary table 2). Study participants came from 31 different countries with highest numbers from the UK (n=88; 21%), Germany (n=76; 18%), USA (n=64; 15%), and Switzerland (n=31; 7%). Participants who completed the special questionnaire on vaccinations were slightly older, more often female, and more often from European countries than those who did not complete the special questionnaire (supplementary table 2).

Vaccination uptake and willingness was high in the study population. Among the 287 adults, 263 (92%) had already been vaccinated, 3 (1%) had an appointment, 7 (2%) planned to get vaccinated, 7 (2%) were hesitant to get vaccinated, and 7 (2%) did not plan to get vaccinated (table 1). People who were unsure or unwilling to get vaccinated more often came from Germany, Switzerland, and other European countries, but we found no difference in age and sex (supplementary table 3). Among the 41 adolescent study participants, 17 (41%) had already been vaccinated and among children below 12 years, 2 of 95 were vaccinated. Five adolescents (12%) and 8 children (8%) did not plan to get vaccinated, even if the vaccine would be approved for persons their age. Among participants who had been vaccinated, most had received Pfizer and BioNTec (48%) followed by AstraZeneca (22%), Moderna (15%), and other (15%).

**Table 1:**
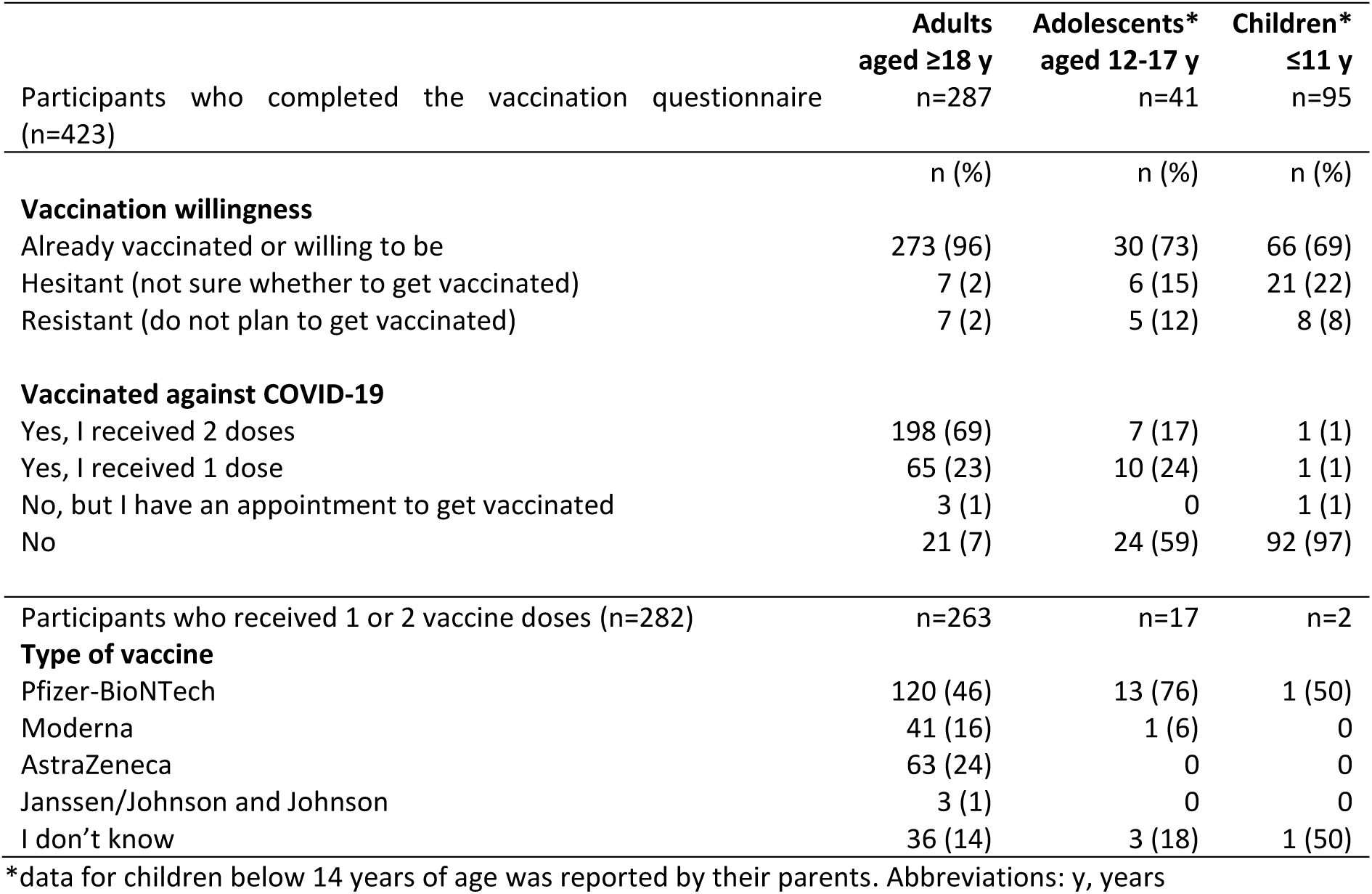
Self-reported vaccination willingness, vaccination uptake, and vaccine type among people with primary ciliary dyskinesia by age (N=423) (COVID-PCD study, May 2021)

|  | **Adults**<br>**aged ≥18 y**<br>**n=287** | **Adolescents\***<br>**aged 12-17 y**<br>**n=41** | **Children\***<br>**≤11 y**<br>**n=95** |
| --- | --- | --- | --- |
| Participants who completed the vaccination questionnaire (n=423) |  |  |  |
|  | n (%) | n (%) | n (%) |
| **Vaccination willingness** |  |  |  |
| Already vaccinated or willing to be | 273 (96) | 30 (73) | 66 (69) |
| Hesitant (not sure whether to get vaccinated) | 7 (2) | 6 (15) | 21 (22) |
| Resistant (do not plan to get vaccinated) | 7 (2) | 5 (12) | 8 (8) |
| **Vaccinated against COVID-19** |  |  |  |
| Yes, I received 2 doses | 198 (69) | 7 (17) | 1 (1) |
| Yes, I received 1 dose | 65 (23) | 10 (24) | 1 (1) |
| No, but I have an appointment to get vaccinated | 3 (1) | 0 | 1 (1) |
| No | 21 (7) | 24 (59) | 92 (97) |
| Participants who received 1 or 2 vaccine doses (n=282) | n=263 | n=17 | n=2 |
| **Type of vaccine** |  |  |  |
| Pfizer-BioNTech | 120 (46) | 13 (76) | 1 (50) |
| Moderna | 41 (16) | 1 (6) | 0 |
| AstraZeneca | 63 (24) | 0 | 0 |
| Janssen/Johnson and Johnson | 3 (1) | 0 | 0 |
| I don't know | 36 (14) | 3 (18) | 1 (50) |

\*data for children below 14 years of age was reported by their parents. Abbreviations: y, years

The most important reasons for getting vaccinated were to protect themselves and others from infection (99%) and to stop the pandemic (95%) (figure 1). Less important reasons to get vaccinated were that the vaccination would enable them to travel and would make activities such as going to the fitness centre possible. Reasons against getting vaccinated were similar among those who did not want to get vaccinated (n=20) and those who were not sure whether to get vaccinated (n=31) (figure 2). In total, 88% of those who were hesitant to get vaccinated reported that they were concerned about side effects and 75% reported concern that the vaccine development had been too rushed. Only few reported reasons relating to disbelief in the effectiveness of the vaccines (8%) or not knowing how to get the vaccine (4%).

**Figure 1:**
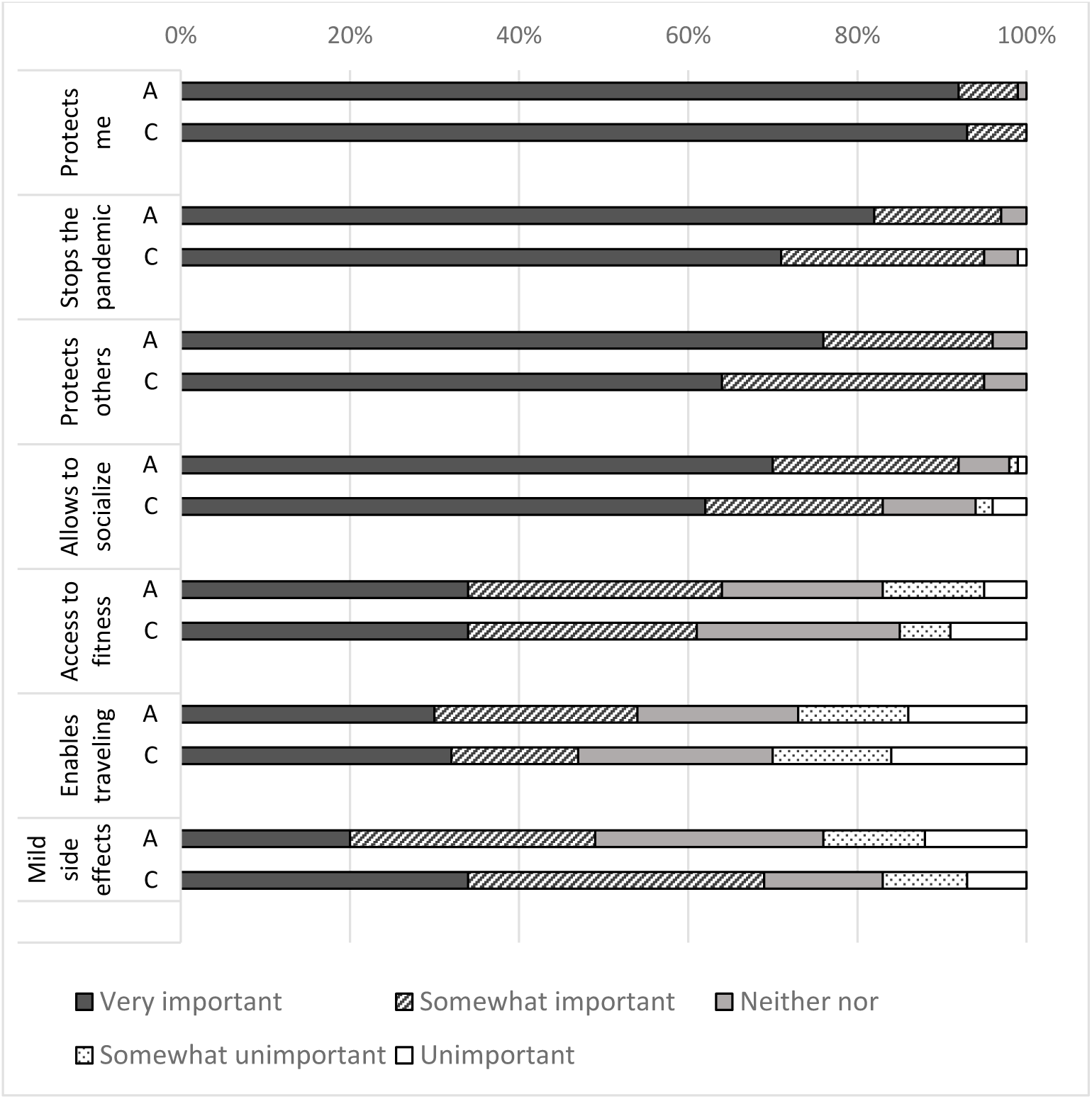
Reasons for getting vaccinated among participants who already got vaccinated or wanted to get vaccinated among adults (n=272) and children and adolescents (n=94). Responses from very important to unimportant. (COVID-PCD study, May 2021) Abbreviations: A = adults aged 18 years or above; C = children and adolescents below 18 years.

**Figure 2:**
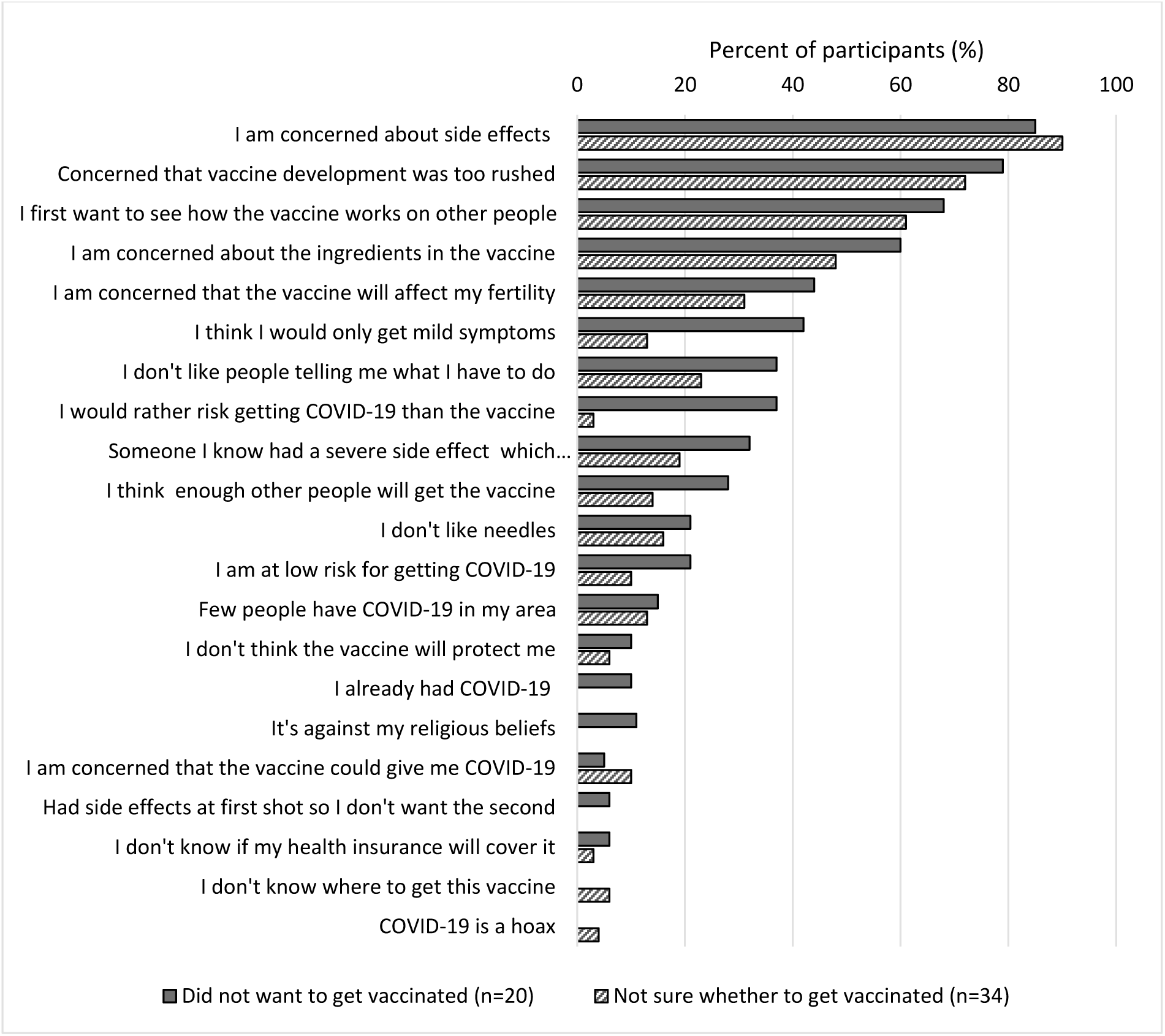
Agreement (somewhat or strongly) with reasons against getting vaccinated among those who did not want to get vaccinated against COVID-19 (n=20) and those who were not sure whether to get vaccinated (n=34). (COVID-PCD study, May 2021) **Note: no difference in responses by age and sex**.

The vaccination uptake and speed of vaccination rollout differed between countries (figure 3). Participants living in the UK were vaccinated faster than in all other countries. By mid-February 2021, 80% of the adults who completed the special vaccination questionnaire had received at least one vaccine injection and by end of June 2021, 100% had received at least one vaccine injection. The second fastest country to vaccinate was USA followed by Switzerland. The country in which the vaccination rollout was slowest was Australia where by June 2021, 80% of participants had received at least one vaccine injection.

**Figure 3:**
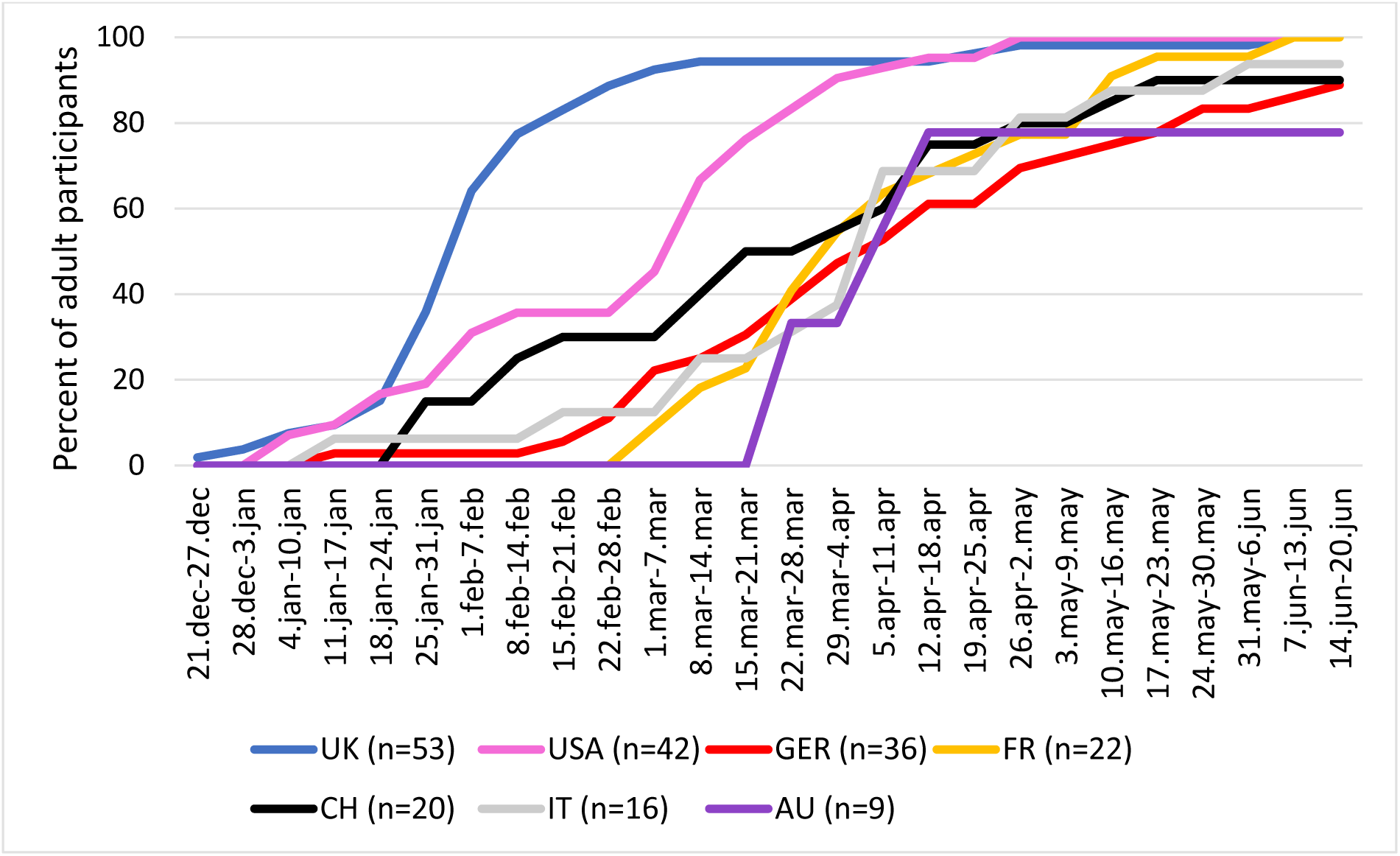
Speed of vaccination uptake among adults in the COVID-PCD in countries with most participants. (COVID-PCD study, May 2021) Abbreviations: UK: United Kingdom, USA: United states of America, GER: Germany, IT: Italy, CH: Switzerland, FR: France, AU: Australia.

Reported side effects after vaccination were common but mild; nobody reported severe side effects. The most common side effect was redness, swelling, or pain around the injection site which was reported by 60% followed by tiredness, headache, aching muscles, and fever (figure 4). Other side effects such as nausea, vomiting, diarrhoea, and stomach ache were rare. Participants more often reported side effects after the second vaccine injection than the first injection, and younger participants more often reported fever, headache, and muscle pain than older participants (Supplementary table 4). Participants who received Moderna or AstraZeneca reported side effects more often than participants who received Pfizer-BioNtech.

**Figure 4:**
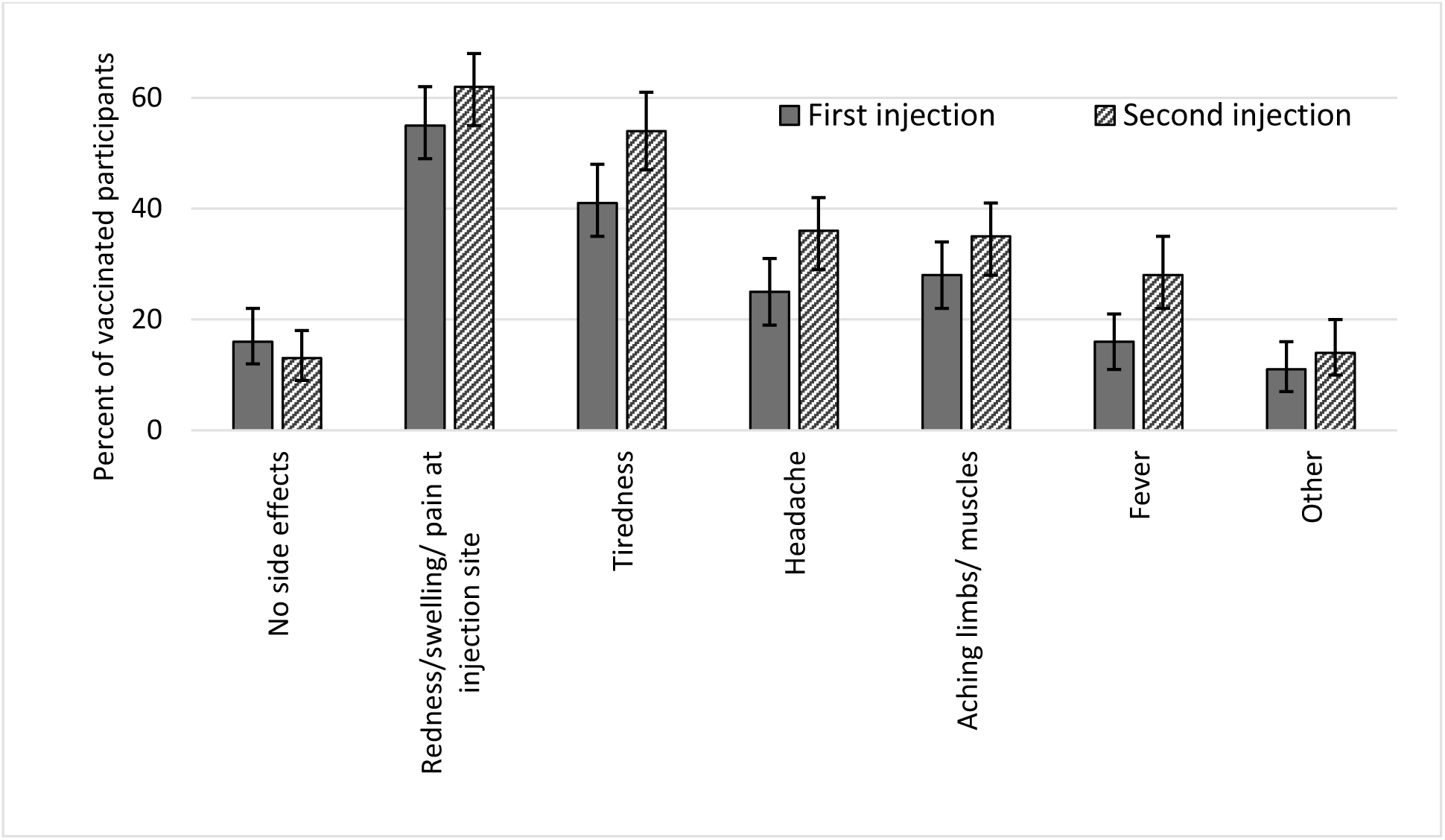
Self-reported side effects after first (n=238) and after second injection (n=214) among vaccinated participants including 95% confidence intervals. (COVID-PCD study, May 2021) Other side effects included: Nausea, vomiting, diarrhea, stomachache, dizziness, chills, breathlessness, cough, congestion, swollen lymph nodes.

Half of the participants changed their social contact behaviour after the first vaccine injection and three quarters after the second vaccine injection (figure 5). After the first injection, participants started to go more often for grocery shopping (reported by 23%) and to meet family and friends (20%). After the second vaccine injection, participants more often saw family and friends (50%), more often went for appointments such as physiotherapy (34%), and more often went shopping (29%) than before.

**Figure 5:**
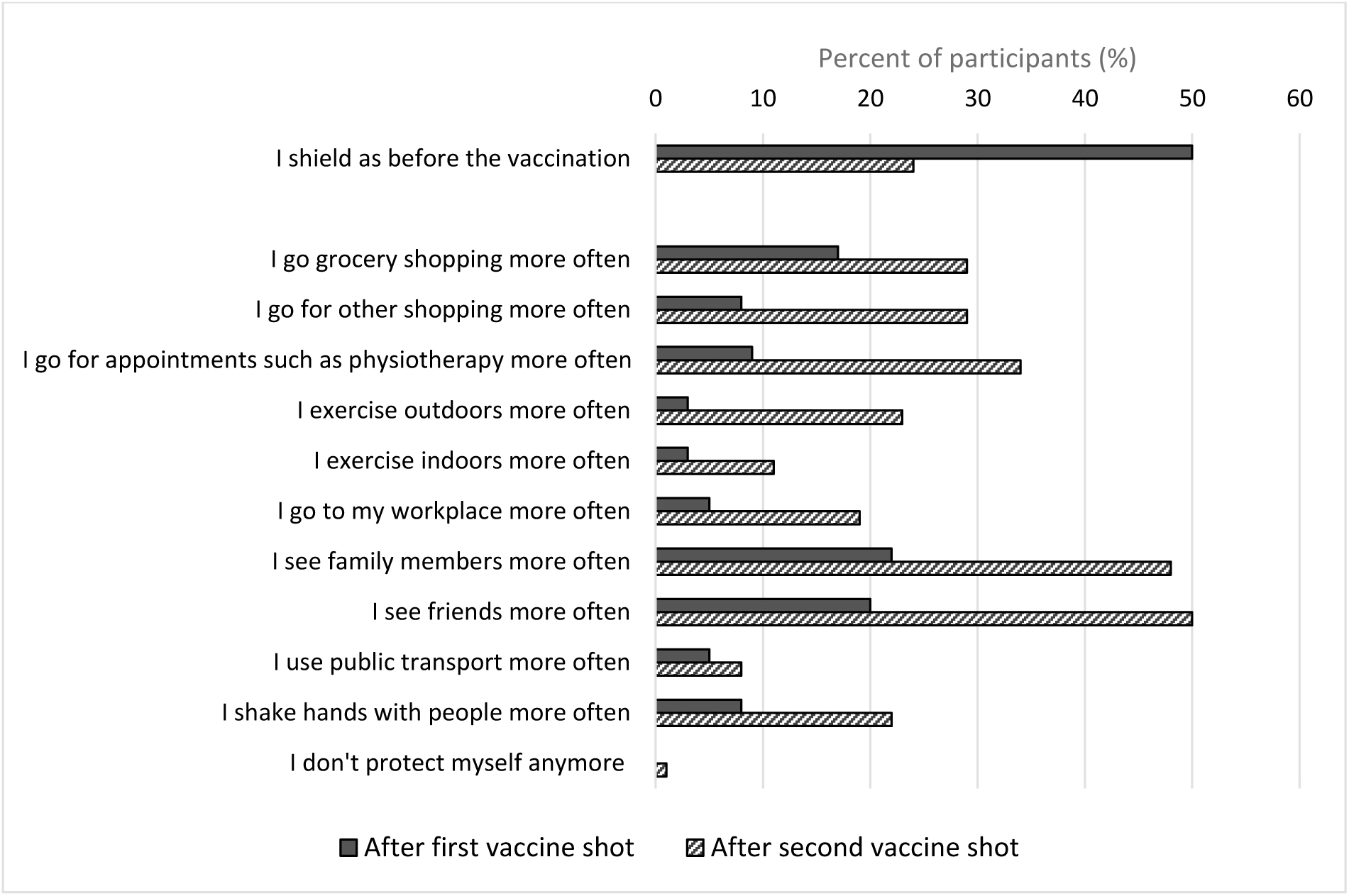
Percent of participants who reported how they changed their social contact behavior after first and second vaccination than before getting vaccinated (N=282). (COVID-PCD study, May 2021)

## Discussion

This international participatory study found that most people with PCD were already vaccinated against COVID-19 or willing to by end of May 2021. Only 2% of adults did not plan to get vaccinated for which the most common reasons were concerns about side effects and concerns that the vaccines had been developed too fast. Side effects were common but mild and were reported more often after the second vaccine injection than after the first, more often by younger participants than older, and more often among participants vaccinated with Moderna or AstraZeneca compared to Pfizer-BioNtech. Half of participants changed their social contact behaviour after the first vaccine injection and mainly started to see family and friends more often.

### Interpretation and comparison with other studies

Vaccination willingness in our study population was high with 96% of adults and 73% of adolescents aged 12-17 years being vaccinated or willing to be by June 2021. Even two children below 12 years had been vaccinated and 69% of parents reported that they were willing to get their child vaccinated if the vaccine would get approved for children. Few studies are published on vaccination uptake and willingness in different countries and most data stem from convenience sampling with high risk of selection bias (30). One study from the UK included more than 2 million unselected adults and found a vaccination willingness of 94% (31). This is as high as we observed in our study population.

However, other studies show lower vaccination willingness in the general population. In a study from the United States including more than 75,000 adults, 80% of the study participants had been vaccinated or were willing to be by end of March 2021 (32). In our study, we found that unwillingness to get vaccinated was more commonly reported in countries such as Switzerland and Germany than in countries such as United Kingdom (supplementary table 3). When looking at whole-country vaccination rates on Our World in Data (8), overall vaccination rates were also higher in the United Kingdom than for example Switzerland. Few studies have reported data on vaccination willingness and uptake in high-risk populations but a study from India among adults with diabetes found a vaccination willingness of 64% (23). Reasons for not getting vaccinated were either fear of side effects, not knowing about the vaccine, or needing to discuss with family members. In our study, nobody reported not to know about the vaccine. PCD support groups around the world have very actively informed members about COVID-19 vaccines and encouraged people with PCD to get vaccinated. This information and encouragement may have contributed to the vaccination uptake and willingness in our population.

We found that side effects were common but mild which has also been observed in the general population. In a large prospective cohort study in more than half a million adults from the UK, local side effects such as pain, swelling, and redness around the injection site were reported by 72% after the first dose of Pfizer-BioNtech and by 59% after first dose of AstraZeneca (33). Systemic side effects such as headache, fatigue, chills and fever were reported by 14% after Pfizer-BioNtech and by 34% after AstraZeneca. Similar findings were seen in two studies among healthcare workers (34, 35). In our population, all side effects were more often reported after the second injection than after the first. We found that systemic side effects were more often reported by participants who received Moderna or AstraZeneca than by people who received Pfizer-BioNtech, which is in line with what was found in the UK study (33). Younger persons were more likely to report side effects than older persons, which has also been shown in other studies and may be explained by a stronger immune responses to the vaccine in younger people (36). Others reported that women were more likely to experience side effect from the vaccines (33, 35). We did not see this. In summary, our study does not suggest that people with PCD have more or worse side effects from the COVID-19 vaccines than the general population.

We found that half of the study participants changed their social behaviour after receiving the second injection of vaccine. Participants started to see family and friends more often, but only very few (1% of participants) reported not to protect themselves anymore after vaccination. There is little published data from the general population on change in social contact behaviour after vaccination but in a study from the UK among 23.287 adults, the authors found no evidence that people did not keep social distancing rules after vaccination (37). In our study, we did not ask whether participants kept social distancing when going out or seeing friends and family but around 20% said that they more often shake hands with people after getting vaccinated. However, we previously found that people with PCD are very careful in protecting themselves against COVID-19 by avoiding many public places and always wearing facemasks (38).

### Strengths and limitations

A major strength of this study is the large sample size of people with PCD from all over the world. PCD is a rare disease, and it can be difficult to recruit participants for research studies. COVID-PCD is a participatory study that was initiated, designed, and tested in collaboration with people who have PCD. This helped to boost study participation. The PCD support groups advertised the special questionnaire on COVID-19 vaccinations via social media and email networks before it was sent out, which may have positively affected the response rate. The questionnaire was completed in five languages which ensured that few people were restricted by language barriers. A limitation is that the study population may not be representative for all people with PCD but mainly those who are in contact with a patient support group. It is difficult to ascertain representativeness of the study population as we have no information about people with PCD who do not participate in the COVID-PCD study. When we compare those who completed the special questionnaire on vaccinations (61%) to those who did not (41%), we see that non-responders were slightly younger than those who responded and more often came from non-European countries (supplementary table 2). It is possible that respondents were more vaccine-willing than non-responders, which would have led to an overestimate of the vaccine uptake. Another limitation is that the sample size, although large for a rare disease, has limited power to detect rare side effects of vaccination.

### Conclusions

In summary, this study found that vaccination willingness and uptake was high among people with PCD which might reflect the extraordinary effort taken by PCD support groups to inform people with PCD about the advantages of vaccinations. No severe side effects were reported. Clear and specific public information about COVID-19 vaccine safety is important for a high vaccine uptake and thus better protection against COVID-19.

## Data Availability

COVID-PCD data can be made available on reasonable request by contacting Claudia Kuehni by.

## Acknowledgements

We thank all participants and their families, and we thank the PCD support groups and physicians who have advertised the study. We thank our collaborators who helped set up the COVID-PCD study: Cristina Ardura, Helena Koppe, Leonie Daria Schreck, Dominique Rubi, University of Bern. We thank Vincenzo Miranda and Federica Annunziata, Federico ll University, Naples, Italy, for helping to translate the study questionnaires to Italian.

## Author contributions

CE Kuehni, ESL Pedersen, F Copland, M Manion, A Harris, JS Lucas, and M Goutaki designed the study. ESL Pedersen, CM Mallet, YT Lam, M Goutaki, and CE Kuehni build the database and collected data. ESL Pedersen drafted the manuscript. All authors contributed to iterations and approved the final version.

## Institutional Review Board Statement

The Bern Cantonal Ethics Committee (Kantonale Ethikkomission Bern) has approved this study (Study ID: 2020-00830) and the research was performed in accordance with the Declaration of Helsinki.

## Data Availability Statement

COVID-PCD data can be made available on reasonable request by contacting Claudia Kuehni by.

## Supplementary information

**Supplementary table 1:**
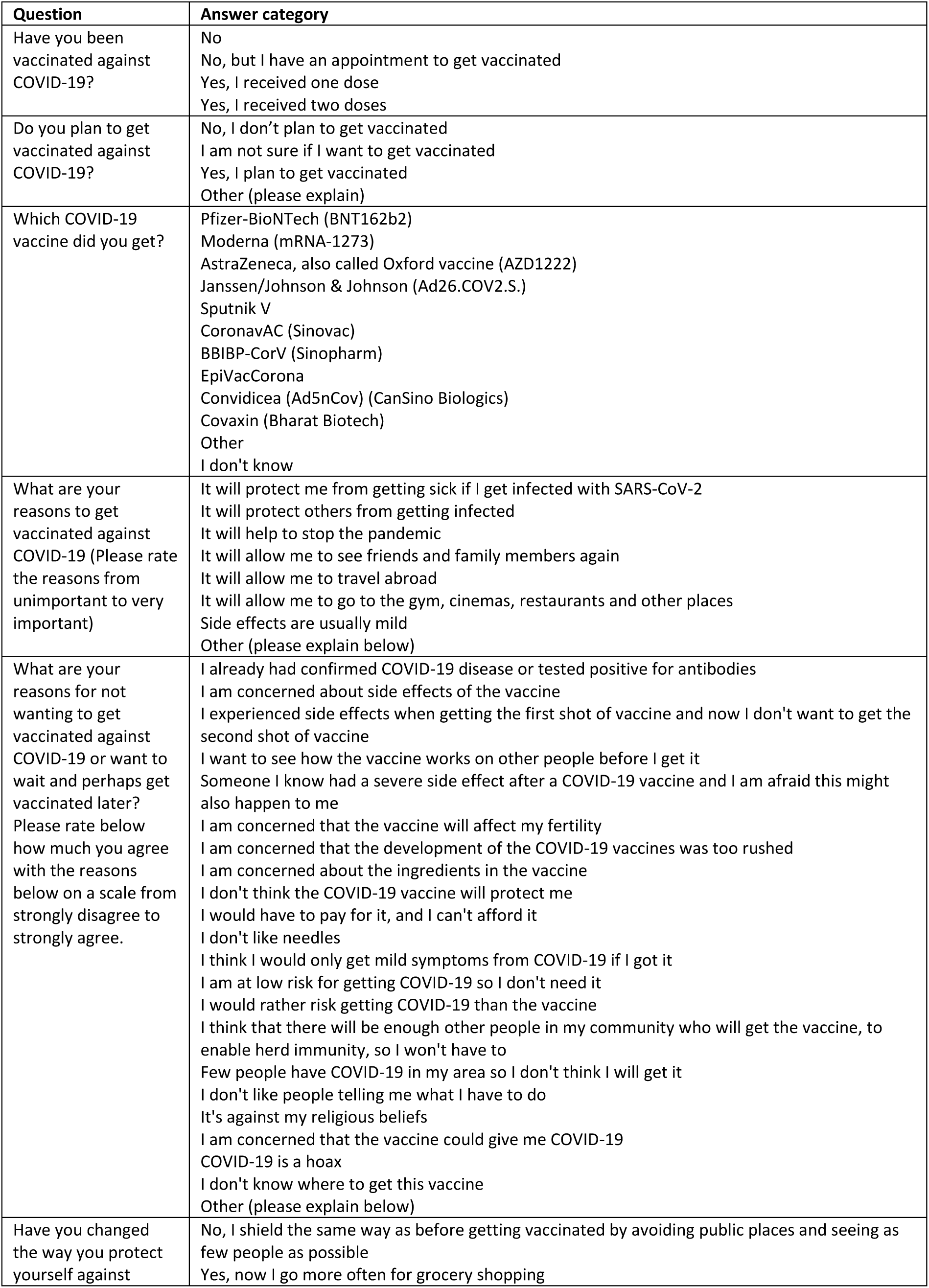
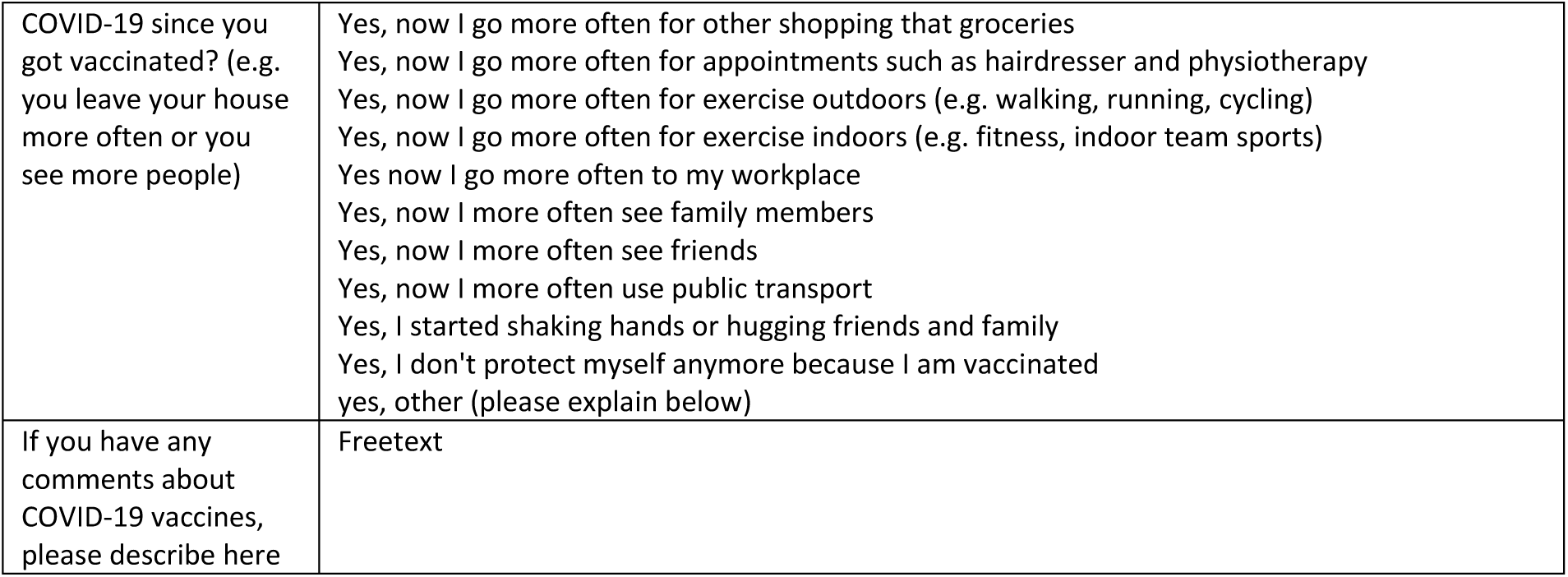
Formulation of questions and answers from the special questionnaire on COVID-19 vaccinations sent to participants in May 2021. (COVID-PCD study, May 2021)

**Supplementary table 2:** Characteristics people with primary ciliary dyskinesia who completed the special questionnaire on COVID-19 vaccinations and those who did not. (COVID-PCD study, May 2021)

|  | Completed<br>vaccination<br>questionnaire | Did not complete<br>vaccination<br>questionnaire |
| --- | --- | --- |
|  | N=423 | N=266 |
|  | n (%) | n (%) |
| **Age, median (range)** | 30 (1-85) | 24 (1-69) |
| **Age groups** |  |  |
| 0-12 y | 95 (22) | 75 (28) |
| 13-17 y | 41 (10) | 24 (9) |
| 18 y or above | 287 (68) | 170 (63) |
| **Sex** |  |  |
| Female | 261 (62) | 152 (57) |
| **Country of residence** |  |  |
| United Kingdom | 88 (21) | 54 (20) |
| USA | 64 (15) | 59 (22) |
| Germany | 76 (18) | 19 (7) |
| Italy | 24 (6) | 22 (8) |
| Switzerland | 31 (7) | 16 (6) |
| France | 23 (5) | 16 (6) |
| Australia | 14 (3) | 13 (5) |
| Other European countries | 81 (19) | 36 (14) |
| Other non-European countries | 22 (5) | 31 (12) |

**Supplementary table 3:** Demographic characteristics of adults with primary ciliary dyskinesia who were already vaccinated or willing to be compared to adults who were unwilling to get vaccinated. (COVID-PCD study, May 2021)

|  | **Vaccinated or**<br>**willing to be** | **Unsure or**<br>**unwilling to get**<br>**vaccinated** | **p-value** |
| --- | --- | --- | --- |
|  | N=262 | N=21 |  |
|  | n (%) | n (%) |  |
| **Age, median (range)** | 41 (30-52) | 37 (25-51) | 0.210* |
| **Sex** |  |  |  |
| Female | 178 (69) | 15 (71) | 0.506# |
| **Country of residence** |  |  | 0.023 |
| United Kingdom | 56 (21) | 2 (10) |  |
| USA | 42 (16) | 1 (5) |  |
| Germany | 37 (14) | 5 (24) |  |
| Italy | 15 (6) | 1 (5) |  |
| Switzerland | 16 (6) | 4 (19) |  |
| France | 19 (7) | 0 |  |
| Australia | 6 (2) | 3 (14) |  |
| Other European countries | 55 (21) | 4 (19) |  |
| Other non-European countries | 16 (6) | 1 (5) |  |

\*Wilcoxon-Mann-Whitney-Test #Fisher's exact

**Supplementary figure 1:**
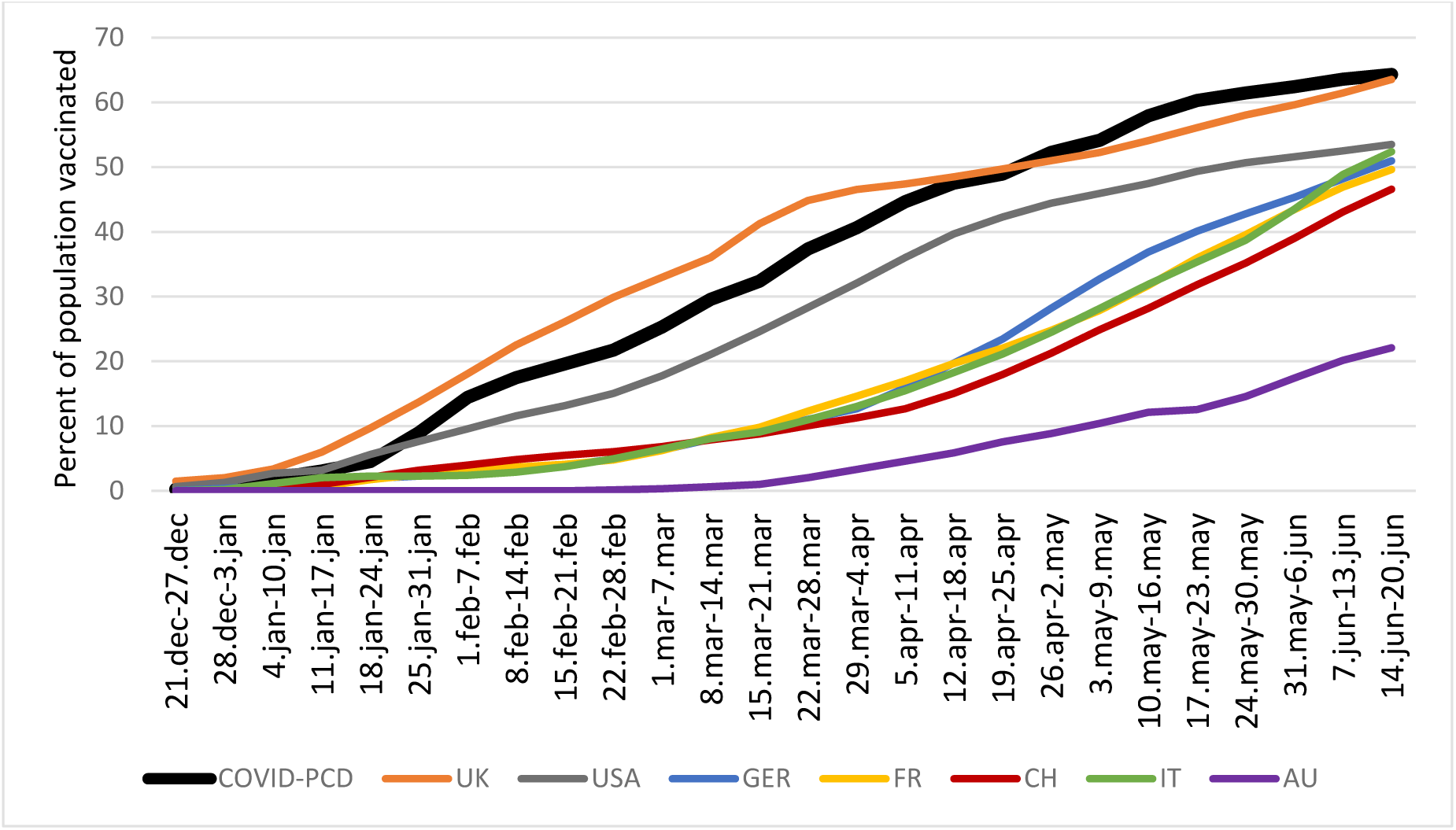
Cumulative percent of COVID-PCD study participants vaccinated (black thick line), compared with nation-wide data from selected countries (colored lines, data source: https://github.com/owid/covid-19-data/tree/master/public/data/vaccinations/country_data, downloaded 24.08.2021) Nation-wide data shown for countries from which at least 15 people participated in the COVID-PCD study by August 2021. Abbreviations: UK: United Kingdom, USA: United states of America, GER: Germany, IT: Italy, CH: Switzerland, FR: France, AU: Australia.

**Supplementary table 4:** Odds ratios of reporting side effects after COVID-19 vaccination adjusted for age, sex, first or second injection, and type of vaccine (N=266). (COVID-PCD study, May 2021)

|  | OR | Lower | Upper | p-value |
| --- | --- | --- | --- | --- |
| **No side effects** |  |  |  |  |
| Age (continuous, per year increase) | 0.98 | 0.95 | 1.00 | 0.126 |
| Sex (female vs. male) | 1.80 | 0.72 | 4.50 | 0.208 |
| Injection (second vs. first) | 1.57 | 0.79 | 3.11 | 0.198 |
| Vaccine (ref: Pfizer-BioNtech) |  |  |  |  |
| Moderna | 2.34 | 0.61 | 8.97 | 0.215 |
| AstraZeneca | 0.89 | 0.33 | 2.42 | 0.818 |
| **Local pain/swelling** |  |  |  |  |
| Age (continuous, per year increase) | 0.99 | 0.96 | 1.01 | 0.320 |
| Sex (female vs. male) | 1.58 | 0.72 | 3.49 | 0.249 |
| Injection (second vs. first) | 1.45 | 0.86 | 2.45 | 0.168 |
| Vaccine (ref: Pfizer-BioNtech) |  |  |  |  |
| Moderna | 3.32 | 1.11 | 9.92 | 0.032 |
| AstraZeneca | 0.54 | 0.22 | 1.30 | 0.167 |
| **Tiredness** |  |  |  |  |
| Age (continuous, per year increase) | 0.99 | 0.97 | 1.01 | 0.374 |
| Sex (female vs. male) | 1.54 | 0.81 | 2.93 | 0.190 |
| Injection (second vs. first) | 2.19 | 1.33 | 3.61 | 0.002 |
| Vaccine (ref: Pfizer-BioNtech) |  |  |  |  |
| Moderna | 2.95 | 1.23 | 7.09 | 0.016 |
| AstraZeneca | 1.12 | 0.55 | 2.29 | 0.751 |
| **Fever** |  |  |  |  |
| Age (continuous, per year increase) | 0.97 | 0.95 | 0.99 | 0.015 |
| Sex (female vs. male) | 0.90 | 0.43 | 1.90 | 0.791 |
| Injection (second vs. first) | 3.12 | 1.64 | 5.94 | 0.001 |
| Vaccine (ref: Pfizer-BioNtech) |  |  |  |  |
| Moderna | 5.64 | 2.06 | 15.42 | 0.001 |
| AstraZeneca | 7.57 | 2.93 | 19.53 | <0.001 |
| **Headache** |  |  |  |  |
| Age (continuous, per year increase) | 0.98 | 0.96 | 1.00 | 0.052 |
| Sex (female vs. male) | 1.73 | 0.83 | 3.62 | 0.147 |
| Injection (second vs. first) | 2.31 | 1.33 | 4.00 | 0.003 |
| Vaccine (ref: Pfizer-BioNtech) |  |  |  |  |
| Moderna | 3.15 | 1.21 | 8.20 | 0.019 |
| AstraZeneca | 6.30 | 2.65 | 15.00 | <0.001 |
| **Muscle/bon pain (not around injection site)** |  |  |  |  |
| Age (continuous, year increase) | 0.97 | 0.95 | 0.99 | 0.003 |
| Sex (female vs. male) | 0.79 | 0.45 | 1.38 | 0.403 |
| Injection (second vs. first) | 1.59 | 0.99 | 2.56 | 0.057 |
| Vaccine (ref: Pfizer-BioNtech) |  |  |  |  |
| Moderna | 2.34 | 1.11 | 4.96 | 0.026 |
| AstraZeneca | 4.73 | 2.34 | 9.53 | <0.001 |
| **Other** |  |  |  |  |
| Age (continuous, per year increase) | 1.01 | 0.97 | 1.04 | 0.751 |
| Sex (female vs. male) | 1.34 | 0.45 | 4.01 | 0.604 |
| Injection (second vs. first) | 0.68 | 0.78 | 3.61 | 0.183 |
| Vaccine (ref: Pfizer-BioNtech) |  |  |  |  |
| Moderna | 3.93 | 0.95 | 16.19 | 0.058 |
| AstraZeneca | 4.22 | 1.24 | 14.38 | 0.021 |

